# A randomized controlled trial comparing epinephrine and dexamethasone to placebo in the treatment of infants with bronchiolitis: the Bronchiolitis in Infants Placebo versus Epinephrine and Dexamethasone (BIPED) study protocol

**DOI:** 10.1101/2024.12.05.24318262

**Authors:** Amy C. Plint, Anna Heath, Tremaine Rowe, Kristina I. Vogel, Natasha Wills-Ibarra, Sharon O’Brien, Meredith L. Borland, David W. Johnson, Joseph J. Zorc, Petros Pechlivanoglou, Suzanne Schuh, Medhawani Rao, Megan Bonisch, Simon S. Craig, Serge Gouin, Amit Kochar, Graham C. Thompson, Chris Lash, Andrew Dixon, Scott Sawyer, Gary Joubert, Ed Oakley, Martin Offringa, Terry P. Klassen, Stuart R. Dalziel

## Abstract

**Background:** Bronchiolitis exerts a significant burden of illness on infants worldwide predominantly due to need for hospitalization. Currently only supportive care is advised by national guidelines for infants with bronchiolitis. There is evidence that treating infants with bronchiolitis with a combination of inhaled epinephrine and dexamethasone may reduce hospital admissions. Synergy between beta-agonists and corticosteroids is recognized in asthma management and basic science literature demonstrates that co-administration of these medications enhances each other’s effectiveness.

**Objective:** To determine if infants with bronchiolitis treated with inhaled epinephrine (delivered by metered dose inhaler with spacer or nebulizer) in the emergency department and a 2-day course of oral dexamethasone have fewer hospitalizations (due to bronchiolitis) over 7 days compared to infants treated with placebo.

**Design:** The BIPED study (Bronchiolitis in Infants Epinephrine versus Dexamethasone and Placebo) is a randomized, placebo-controlled, observer, investigator, clinician, and patient blinded superiority clinical trial being conducted in 12 emergency departments across three countries (Canada, New Zealand, and Australia). We will recruit 864 infants between 60 days and 12 months of age with bronchiolitis to receive either (1) two inhaled epinephrine treatments (3 mg via nebulizer or 625 mcg via metered dose inhaler with spacer) 30 minutes apart and a simultaneous dose of oral dexamethasone (0.6 mg/kg, maximum 10 mg) in the emergency department with the dexamethasone repeated at 24 hours or (2) inhaled placebo and oral placebo. The primary outcome is hospital admission for bronchiolitis within 7 days (168 hours) of enrolment. Secondary outcomes include hospital admission during enrolment, and all cause hospital admissions and healthcare provider visits within 21 days of enrolment. We will use a Bayesian approach for data analysis and inference.

**Discussion:** Given the healthcare burden of bronchiolitis, there is urgent need for a trial to confirm if combination therapy with epinephrine and dexamethasone is effective.

**Trial registration:** ClinicalTrials.gov: NCT03567473. Registered on 2018-06-13.

**Protocol version:** CTO 1423, dated 20 January 2023

**Sponsor-Investigator:** Dr. Amy C Plint, Children’s Hospital of Eastern Ontario, Ottawa, Ontario, Canada

**Secondary Sponsor:** Children’s Hospital of Eastern Ontario Research Institute, Ottawa, Ontario, Canada

## INTRODUCTION

Bronchiolitis is the leading cause of infant hospitalization in North America and accounts for a substantial burden of illness and health-related expenditures[1–5].In the United States, it is estimated that there are 100,000 hospitalizations annually for bronchiolitis at a cost of $1.73 billion[5]. In Canada 35 per 1,000 infants younger than one year of age are hospitalized annually with an estimated cost of more than $23 million[2, 6]. These high admission rates persist every bronchiolitis season, despite clinicians’ best efforts to prevent hospitalization, where possible.

To date, two large multicenter trials have generated key evidence in the treatment of bronchiolitis with the aim of reducing hospitalizations. The first trial, conducted in the US through the Pediatric Emergency Care Applied Research Network (PECARN), found that treatment of infants with a single dose of oral dexamethasone did not reduce hospitalizations[7]. The second trial (Canadian Bronchiolitis Epinephrine Steroid Trial – CanBEST), conducted in Canada through Pediatric Emergency Research Canada (PERC), confirmed that oral dexamethasone did not reduce hospitalization but found that that treating infants with co-administered nebulized epinephrine and dexamethasone had reduced symptoms sufficiently to decrease hospitalization by one third[8]. Given that this was an unanticipated finding, CanBEST was not adequately powered to definitively test for synergy between these drugs. While other clinical and basic science studies[9–18] support CanBEST’s findings, the suggested synergy from combined treatment raised considerable controversy with divergent views[19–23]. Further, a survey of Canadian pediatric emergency physicians revealed that combined dexamethasone and epinephrine therapy is not routinely used, with the majority of physicians demanding another trial to confirm that combined therapy is safe and effective before they will their change practice[24]. Most physicians surveyed indicated a strong preference to use a shorter course of dexamethasone, similar to croup management. This desire to use a shorter course of dexamethasone is supported by the CanBEST study seeing the benefit of combined epinephrine and dexamethasone therapy on hospitalizations being present within the first few days and with several previous small studies also providing support for such dosing[9–11, 25]. Currently no published guidelines for bronchiolitis endorse the use of combined epinephrine and dexamethasone therapy in the treatment of bronchiolitis, with most citing the need for further evidence[26–28] #199]. Only supportive care (e.g., fluids and oxygen) is recommended[29].

Given the large health care burden of bronchiolitis, the preliminary evidence that the combination of epinephrine and dexamethasone can substantially reduce hospitalizations and the divergent current opinion, there is an urgent need for a trial to confirm if combination therapy with epinephrine and dexamethasone is effective in comparison to the current standard of supportive care, only. The objective of our study is to determine if infants with bronchiolitis who are treated with inhaled epinephrine (delivered by MDI-s or nebulizer) in the ED and a 2-day course of oral dexamethasone have fewer hospitalizations (due to bronchiolitis) over 7 days compared to infants treated with placebo, without relevant drug related side effects. The patient-based outcomes assessed in this trial – hospitalization rates, symptoms, costs of care, and risk of adverse events - will be of wide interest to caregivers, clinicians, and health care administrators alike.

## METHODS AND ANALYSIS

### Hypothesis

Infants with bronchiolitis treated with inhaled epinephrine in the ED and a 2-day course of oral dexamethasone will have fewer hospitalizations over 7 days compared to infants treated with placebo

### Trial design

BIPED is a phase III, 12-centre, randomized, controlled, observer, investigator, clinician, and participant blinded superiority clinical trial with two parallel groups. Randomization will be performed as block randomization with 1:1 allocation. The primary endpoint is admission to hospital due to bronchiolitis within 7 days (168 hours) of enrolment.

### Study setting

Recruitment will occur at six pediatric academic EDs across Canada (Centre Hospitalier Universitaire Sainte-Justine, Montreal; Children’s Hospital of Eastern Ontario, Ottawa; Children’s Hospital London Health Sciences Centre, London; Children’s Hospital, Winnipeg; Alberta Children’s Hospital, Calgary; Stollery Children’s Hospital, Edmonton), three academic hospitals in New Zealand (Starship Children’s Hospital, Auckland; Middlemore (Kidz First) Hospital, Auckland; Waikato Hospital, Hamilton) and three pediatric academic hospitals in Australia (Women and Children’s Hospital, Adelaide, South Australia; Monash Medical Centre, Melbourne, VIC; Perth Children’s Hospital, Perth, WA). All participating EDs in Canada are members of the PERC research collaboration and all participating EDs in New Zealand and Australia are members of the Pediatric Research in the Emergency Department International Collaborative (PREDICT). The annual ED census for the participating centres ranges from 20,000 to 90,000, each site treats hundreds of bronchiolitis cases annually. Research Ethics Board (REB) approval has been granted by the board of record at each participating site.

### Eligibility Criteria

#### Inclusion criteria

All the following criteria must be met for inclusion in the study.

1. Aged 60 days to less than 12 months of age
2. Presenting to the ED with an episode of bronchiolitis. Bronchiolitis will be defined as an episode of wheezing or crackles associated with signs of an upper respiratory tract infection (e.g., cough, coryza, nasal congestion) during the period deemed to be the season for respiratory syncytial virus (RSV) bronchiolitis (traditionally approximately December to April in Northern Hemisphere and June to October in Southern Hemisphere). We have chosen not to define bronchiolitis as the first episode of wheezing or crackles to better reflect the clinical guidelines and clinical practice internationally[29].

#### Exclusion criteria

Children who meet any of the following criteria will be excluded from participation in the study.

1. Respiratory distress assessment instrument (RDAI) score of ≤ 3. The RDAI, has good interobserver reliability, rates wheezing and respiratory retractions on a scale from 0 to 17, with higher scores indicating more severe illness; a score below 4 indicates very mild illness, and a score above 15 very severe illness[30, 31].
2. Previously known chronic disease that may affect cardiopulmonary status of the patient (e.g., bronchopulmonary dysplasia currently receiving oxygen, cystic fibrosis, congenital heart disease and immune deficiency).
3. Severe respiratory distress evidenced by a sustained pulse rate >200 beats/min, a sustained respiratory rate >80 breaths/min, profound lethargy or requiring resuscitation room care.
4. Presenting with symptoms of apnea prior to enrolment.
5. Treatment with oral, inhaled, or intravenous corticosteroids within the last 1 week.
6. History of adverse reaction to glucocorticoids.
7. Treatment with any beta-agonists (salbutamol/albuterol or epinephrine/adrenaline) in the ED prior to study enrolment.
8. Presence of varicella or recent (less than 3 weeks) close contact without a history of prior infection.
9. Insurmountable language barrier.
10. Any child born at less than 37 weeks gestation who is younger than 60 days corrected age.
11. Previous enrolment in the trial.
12. Unavailability for follow-up period.
13. Certain admission to hospital.

### Interventions

Participants will be randomized to either (a) active intervention: oral dexamethasone and inhaled epinephrine delivered by MDI-s (Primatene MIST ^R^) or by nebulization (1:1000 epinephrine), or (b) control intervention: oral placebo (OraBlend^R^ or OraMix SF^R^ in Canada and a compounded oral placebo in New Zealand and Australia) and inhaled placebo by MDI-s or nebulized saline. Currently, only supportive care (e.g., fluids and oxygen) is recommended for the management of bronchiolitis and thus, the selection of placebo as the control arm for this trial is appropriate[26, 32, 33]. Combined therapy with oral dexamethasone and nebulized epinephrine has shown promise in reducing admission to hospital for infants with bronchiolitis[8] and as such will form the active intervention arm.

#### Active intervention arm

Participants randomized to the active intervention arm will receive oral dexamethasone (0.6 mg/kg; maximum dose 10 mg) in the ED immediately prior (within 15 minutes) to their first epinephrine treatment. If a participant vomits the oral dexamethasone within 15 minutes of their dose, the dose will be repeated. If they vomit a second time, the dose will not be repeated. Following the oral dexamethasone, they will receive two treatments of inhaled epinephrine 30 minutes apart (+/− 15 minutes). Each treatment of inhaled epinephrine will be either 625 mcg of epinephrine via MDI-s (5 actuations of Primatene MIST^R^ for each treatment) or 3 mL of 1:1000 epinephrine by nebulizer. The spacer used will be the AeroChamber Plus Flow-Vu^R^ with facemask, size small). If the study site chose to utilize nebulizer for study medication administration, the epinephrine will be administered by a Medline Hudson Micromist Nebulizer^R^ with face mask. Twenty-four hours (range 22 to 40 hours) after their first dose of dexamethasone in the ED they will receive a second dose of dexamethasone (0.6 mg/kg; maximum dose 10 mg) either at home or in hospital (if admitted to hospital). This second dose of dexamethasone will not be repeated if vomited.

#### Control intervention arm

Participants in the control arm will be managed in the same manner as those in the active intervention arm but will receive 0.6 mL/kg (maximum 10 mL) of the oral placebo in the ED and 24 hours after their first dose. Inhaled placebo treatment will consist of 5 actuations from a placebo MDI-s identical in appearance to the active product (and delivered as described above for the active treatment) or 3 mL of normal saline by nebulization (delivered by the Hudson Micromist Nebulizer^R^).

#### Rationale for study interventions dosing and regime

The epinephrine dose was chosen based on the CanBEST trial and other studies utilizing epinephrine in bronchiolitis[8, 34–38]. In CanBEST, participants received two doses of 3 mL of 1:1000 epinephrine (3 mg per dose) by nebulizer and for participants receiving a nebulized intervention we will use this dose. Evidence from the asthma literature suggests that 5 mg of salbutamol by nebulizer is equivalent to 1000 mcg by MDI (given as 10 actuations of 100 mcg/actuation)[39–41]. Extrapolating from this evidence, 3 mg of epinephrine by nebulizer is equivalent to 625 mcg epinephrine delivered by MDI. Primatene MIST^R^ delivers 125 mcg of epinephrine per actuation and thus we will use 5 actuations to deliver 625 mcg per treatment.

While the CanBEST trial used 6 days of dexamethasone, all the benefit of combined epinephrine/dexamethasone therapy occurred in the first few days[8]. Given this observation from CanBEST and knowing that dexamethasone exerts effects for up to 72 hours[42], we have shortened the course to 2 days. We have also reduced the initial ED dose from 1.0 mg/kg to 0.6 mg/kg, knowing that 0.6 mg/kg dosing will achieve plasma and tissue concentrations sufficiently high to achieve maximal synergistic upregulation of mRNA production of glucocorticosteroid-inducible anti-inflammatory genes[17, 43, 44] and is consistent with the higher-end dose used in croup. Available basic science evidence suggests that close timing between administration of glucorticoids and beta-agonists is critical to up-regulating anti-inflammatory gene expression[45].

#### Discontinuing or modifying allocated interventions

No dose modification of the study medications will be permitted within the protocol. The allocated intervention may be discontinued by the site qualified investigator (QI) or attending ED clinician if there are clinical concerns that may be related to study interventions or at caregiver request. Caregivers may also choose, at their discretion, to not give the second oral dose of medications at home.

#### Study medication compliance

Research nurses will administer the inhaled medications and oral medications in the ED minimizing compliance concerns with these medications. To minimize any compliance problems with the remaining oral medication dose, participants will be sent home with either their pre-measured dose of dexamethasone or placebo in an oral syringe or with an oral syringe and bottle of dexamethasone or placebo with clear instructions of dose to administer. Dexamethasone is palatable, easily administered and well tolerated. Caregivers will be asked to administer the dexamethasone 24 hours after the ED dexamethasone dose but will be advised they can administer it within a 22 to 40-hour window due to the infant’s sleep or feeding schedule. Compliance with home medication administration will be ascertained during telephone/email follow-up (including date and time of administration, dose given, individual administering the intervention, and method of discarding of the empty syringe).

#### Concomitant care and medications

Within the ED, patients may receive bronchodilator co-interventions (epinephrine or salbutamol) after administration of study medications at the discretion of their attending ED clinician. Other co-interventions such as antibiotics and intravenous fluids are allowed at any time during the ED visit. Non-study discharge medications are at the discretion of the attending ED clinician. The participant’s ED medical record will serve as the source document for determining concomitant therapy in the ED and at discharge.

### Randomization, allocation concealment, and blinding

A statistician (not otherwise involved in the study) will produce and maintain a computer-generated permuted-block randomization list, stratified by study site. The block sizes will not be disclosed, to ensure concealment. Participants will be randomly assigned to either the control or active intervention arm with a 1:1 allocation scheme. As respiratory viruses vary throughout the year (and could theoretically influence response to therapy) the use of block randomization will ensure comparable distribution of the patients for each group throughout the season.

At Canadian sites, the randomization list will be sent to each participating site’s research pharmacist. The site research pharmacist will keep the list concealed and use it to create pre-packaged, sequential study kits. For Australian sites, a central research pharmacist at Optima Ovest will receive the randomization list and keep it concealed. The Optima Ovest pharmacy will prepare sequential kits containing the study treatments in sealed, opaque numbered bags and provide these to the participating Australian hospitals. In New Zealand, the Auckland City Hospital research pharmacy will receive the randomization list and keep it concealed. The Optima Ovest pharmacy will also prepare study drugs for the New Zealand sites, but the Auckland City Hospital research pharmacy will prepare the study packages and distribute them to the participating New Zealand sites. The active and placebo drugs will be identical in appearance, volume, weight, smell, and taste. Study drugs will be packaged identically and identified only by a sequential study number. Once participant eligibility has been confirmed by the treating ED clinician and consent for enrolment has been obtained by the research nurse, they will select the next-in-sequence study medication kit. As the study drugs are packaged identically and identified only by a sequential study number, the research nurse who administers the intervention will be unaware of the next group assignment.

All study personnel (including research nurses, coordinators, investigators, data management staff, and statistical team), health care staff providing patient care, and participants/caregivers will be blinded to the study group assignment. Following locking of the database and completion of the main study analysis the study team will be unblinded.

Unblinding may occur if the treating ED clinician, or subsequent inpatient physician, demonstrates that knowledge of treatment allocation will alter clinical care. Unblinding decisions need to be discussed and approved by the site qualified investigator (QI) and sponsor-investigator. In the unlikely event of neither the site QI nor sponsor-investigator being available, the treating ED clinician, or subsequent inpatient physician, may make the decision to unblind. During daytime hours the site pharmacist will reveal the group allocation. Outside of daytime hours, unblinding will occur using a Research Electronic Data Capture (REDCap) database that will require the user to enter a kit number, reason for unblinding and provide details of contact with sponsor-investigator and site QI before unblinding occurs. Only site coordinators and the sponsor-investigator will have access to the REDCap unblinding database. Any access of this database is tracked. The individual accessing the unblinding database will reveal the group allocation to the treating ED clinician, or subsequent inpatient physician, and if required to comply with REB and regulatory reporting requirements to the site QI and sponsor-investigator. The Data Safety Monitoring Board (DSMB) may request unblinding from the statistician holding the master list if they deem necessary to consider the results of the interim safety analysis or to review possible adverse events (AEs).

### Recruitment and data collection

Figure 1 outlines the schedule of activities for the trial. Research nurses will be present in the ED to approach and screen infants for eligibility following site specific REB guidelines regarding approaching patients and families for research studies. ED clinicians (ED physician or ED nurse practitioner) will confirm the participant’s eligibility. The research nurse will obtain written informed consent from the legal guardians of eligible infants.

**Figure 1:**
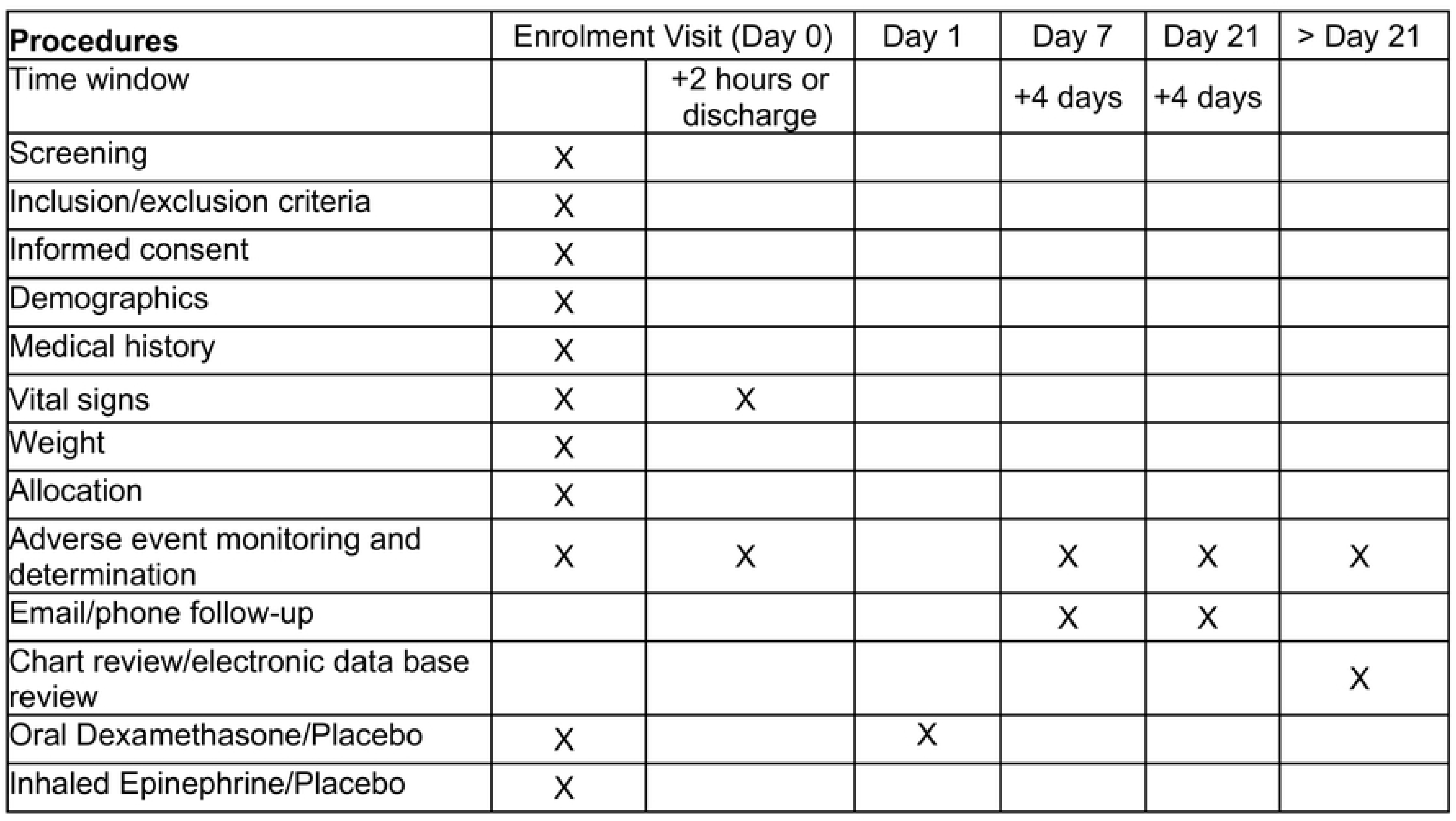
Schedule of trial activities.

After obtaining consent the research nurse will obtain and record on standardized case report forms (CRFs) the participant’s respiratory rate, heart rate and oxygen saturation measured prior to the participant receiving their oral study medication (time 0). Blood pressure will be measured at enrolment when possible. Triage ED temperature measurement will be used as the baseline temperature. The research nurse will then select the next sequential study kit administer the study drug interventions. Information regarding side effects, co-interventions, length of stay and disposition will be recorded throughout the ED intervention period of 2 hours following administration of the oral medication or until discharge (whichever is first). During the ED visit the research nurse will interview the participant’s caregiver to collect demographic details, details regarding current illness, past medical history, and family history. The research nurses will obtain and record all ED based data on standardized CRFs (paper forms in New Zealand and Australia and electronic CRFs (e-CRFS) with direct data entry in Canada).

Two brief standardized surveys will be completed by the participants’ caregiver at day 7 and 21 following enrolment. Caregivers will be given the option of completing the survey by email or telephone. The follow-up surveys will identify subsequent admissions, ED visits, and health care visits, patient symptoms, other medication usage, parental absence from employment and other patient/family costs related to their illness. Adverse outcomes will be also explicitly monitored during survey follow-up (e.g., severe varicella, signs of gastro-intestinal bleeding, serious bacterial infections) and we will also track unsolicited AEs. We will send a second email to those parents/caregivers who select email as the method of follow-up and do not complete the surveys. For families who select telephone as the first method of follow-up, we will call families we cannot reach on the day following their scheduled follow-up day (i.e., day 8 and day 22 as needed). We will make at least 6 attempts over 3 different days to reach families to complete the scheduled follow-up surveys as needed.

A research team member will review each participant’s medical record once the 21-day follow-up period has elapsed to examine return ED visits, admission to hospital, diagnosis for any hospital visit, treatments as well as laboratory results. In the regions served by the participating study hospitals, the majority of infants who will participate in the study would visit the ED or be admitted to the participating study hospitals. All ED visits and admissions reported by the participant’s caregiver will be confirmed by reviewing the medical record review at the study site. If reported ED visit or admission to a study hospital is not confirmed through a medical record review, then no ED visit or admission will have been deemed to occur. For six study sites, ED visits and admissions to nonparticipating hospitals can and will be similarly confirmed through an administrative database record review. All children where the final diagnosis is coded as bronchiolitis will be considered to have visited the ED or been admitted for bronchiolitis. All medical records where the diagnosis is other than bronchiolitis will be reviewed by the site QI to determine reason for ED visit or hospital admission. For study sites where ED visits and admissions to nonparticipating hospital ED visits cannot be confirmed by administrative database record review, the caregiver’s report of and reason for these visits/admissions will be used to determine whether the participant met the study definition of admission. At all sites, data collection following the ED enrolment visit will utilize e-CRFs and direct data entry.

### Outcomes

#### Primary Outcome

The primary outcome will be admission to hospital for bronchiolitis within 7 days (168 hours) of study enrolment. Rates of hospital admission are of interest to clinicians, caregivers, and health system administrators. We have chosen to examine all admissions up to 7 days rather than admissions from the ED, as previous studies have shown that up to 32% of admissions for bronchiolitis occur after initial ED presentation[8]. The attending ED clinician will make the disposition decision based on their clinical judgement as in our previous study[8]. Objective criteria for admission are lacking in clinical practice guidelines, vary widely across institutions, and would be impractical to standardize in the study or generalize to routine practice. While most participants will likely be physically admitted to an inpatient bed, at times patients stay in the ED as “boarded inpatients” due to bed shortages. We will therefore define hospitalization by either: 1) participant being admitted to inpatient ward or 2) a total ED length of stay 12 hours or greater or 3) a combined ED and observation unit stay of 12 hours or greater.

#### Secondary outcomes

a. Admission to hospital for bronchiolitis at the time of the enrolment ED visit;
b. All cause admission to hospital within 21 days following enrolment ED visit;
c. All cause health care provider visits (including ED visits) by day 21 following enrolment ED visit;
d. Health care related costs within the 21 days following enrolment ED visit.

#### Safety outcomes

a. Gastrointestinal bleeding involving either melena or frank blood per rectum (and not attributable to other causes, as determined by the treating physician);
b. Development of serious bacterial infection (defined as meningitis, osteomyelitis, or septicaemia);
c. Severe varicella including arthritis, osteomyelitis, symptomatic hepatitis, pancreatitis, cerebritis, cerebellitis, pneumonitis, glomerulonephritis, disseminated intravascular coagulation, thrombo-cytopenia, prolonged vesicular rash (>3 weeks), fasciitis, septicaemia, ocular complications, orchitis, myocarditis, intensive care admission and death[46];
d. Death.

#### Exploratory Outcomes

a. Admission to hospital for bronchiolitis within 21 days following enrolment ED visit;
b. Admissions to the intensive care unit (ICU) within 21 days following enrolment ED visit for bronchiolitis and requiring intubation or continuous positive airway pressure (CPAP);
c. All cause admission to hospital within 7 days following enrolment ED visit;
d. All cause ED visits within 21 days following enrolment ED visit;
e. Length of stay for the enrolment ED visit (in hours), defined as discharge time minus oral study medication time, for participants discharged at the enrolment ED visit;
f. Length of hospital admission (in hours) for those patients admitted at their enrolment visit (defined as time of hospital discharge minus the time of oral study medication);
g. Resolution of symptoms as documented on a standardized questionnaire during telephone or email at day 7 and 21 days;
h. Out of pocket expenses borne by the family/caregiver within the 21 days following the enrolment ED visit;
i. Age-dependent variation in the efficacy of epinephrine and dexamethasone;
j. Health care utilization (including ambulatory visits, ED visits, hospitalization) for respiratory illness up to 18 years of age;
k. Development of respiratory illnesses such as asthma and recurrent wheezing.

### Sample size

The sample size was determined using he average length criterion (ALC) for Bayesian analysis. The ALC selects the smallest sample size for which the average length of the 95% high density posterior credible interval for the difference in admission rates between placebo and active medication is below 9%. The prior distributions for the probability of hospitalization under placebo and active medication were obtained using expert elicitation[47]. These prior distributions were used both in the posterior distribution and to determine the prior-predictive distribution of the trial data. Using 1,500 simulations from the prior-predictive distribution and 5,000 simulations from the posterior to estimate the average length of the 95% posterior credible intervals, the sample size of 410 participants per arm for the BIPED study fulfils the ALC[47]. We adjust for a 5% loss to follow-up, resulting in a total recruitment of 864 participants, 432 per arm.

### Statistical methods

#### Primary and secondary outcomes

The final trial analysis will use a Bayesian inferential framework[48] based on the posterior probability that the epinephrine and dexamethasone combination is superior to placebo. It will follow the intention-to-treat principle. The epinephrine and dexamethasone combination will be declared superior to placebo if the posterior probability of superiority is above 99%, with simulations confirming a Type 1 error rate of 4.1% with a probability of hospitalization equal to 0.35 and a power of 81% with a target difference of 8%. Due to the likelihood principle, corrections for multiple comparisons will not be used[49]. All estimates for the treatment effect will be reported using 95% high density posterior credible intervals, calculated using R[50] interfacing with Bayesian software such as JAGS, stan or INLA[51, 52].

The primary effectiveness analysis will compute the posterior probability that epinephrine and dexamethasone used together results in a lower probability of hospitalization than placebo. We will use hierarchical Bayesian relative risk regression to adjust for site with an informative prior for the treatment effect, obtained from our expert elicitation exercise[47]. We will also report the mean and 95% high-density posterior credible interval for the absolute risk difference and relative risk across the two treatment groups. We will undertake a secondary analysis for the primary outcome with participants presenting with a first episode of bronchiolitis only. We will undertake this analysis to reflect the variability among researchers and clinicians in whether the definition of bronchiolitis includes only the first episode of wheezing or crackles[7, 8, 53–56]. Apart from the treatment effect, we will use weakly informative priors for the variance and coefficients[57]. Secondary outcomes (a) thru (c) are binary and will be analyzed using relative risk regression, like the primary outcome, with weakly informative priors for all parameters. We will report the probability that active medication is superior to placebo, the mean absolute risk difference and relative risk and 95% credible intervals. For secondary outcome of costs related to health care expenditure, we will undertake a separate formal economic analysis. There are no planned interim analyses of efficacy outcomes[58]. A full statistical analyses plan (SAP) will be published separately prior to data lock and analysis.

#### Safety Outcomes

The comparison of safety outcomes one, two and three across treatment groups will use relative risk regression, like the primary outcome, with weakly informative priors for all parameters. We will report the mean absolute difference, the 95% credible interval and the probability of increased chance of adverse events for active intervention arm. We will also report the number and percentage of observed events for each of the four safety outcomes. All adverse events will be coded using Medical Dictionary for Regulatory Activities (MedDRA)[59] and each event type will be counted at most once for each participant. We will then present the severity and frequency of the adverse event by System Organ Class.

#### Exploratory outcomes

Exploratory outcomes will be analyzed using a Bayesian generalized linear modelling framework. Relevant treatment effect summaries will be provided using the posterior median and corresponding 95% credible intervals. Age dependent effects will be estimated using interaction effects. More details are provided in the SAP.

#### Planned subgroup analyses

We will undertake five planned subgroup analyses in which we will estimate the treatment effect for the primary outcome within each subgroup. These subgroups will include:

1. onset of illness within 48 hours: yes/no (with onset of illness defined as the development of first respiratory symptoms including coryza and cough or the onset of fever) including coryza and cough or fever, within 2 days of enrolment
2. personal history of eczema: yes/no
3. family history of atopy (allergies/eczema/asthma in parents or siblings); yes/no
4. patients stratified by hemisphere of participating hospitals: north/south
5. patients stratified by receipt of nebulized or MDI-spacer medications: yes/no

For each of these subgroup analyses, we will use relative risk regression with main effects for the subgroup assignment and treatment group and an interaction between treatment and subgroup. If the interaction effect has a 99% probability of being greater or less than 0, we will declare that a positive or negative subgroup effect, respectively.

#### Protocol non-adherence and missing data

The primary analysis will be intention-to-treat and include all randomized participants for whom outcome data is available and who do not withdraw consent for the use of their data. We will also undertake a per protocol analysis. The per protocol analysis will include only eligible participants who completed their ED based medications (both doses of inhaled study medication and the dose of oral medication) and their second dose of oral medication, for whom we have outcome data, and who have not withdrawn consent for the use of their data. Descriptive statistics on the proportion of missing data by treatment arm will be provided for the outcomes and the covariates. If the level of missingness is below 5%, we will undertake a complete case analysis. If greater than 5% of the data are missing, we will evaluate if there are any differences between the baseline characteristics for participants with and without missing data. Using this evidence, we will determine whether a complete case analysis is valid. If covariate outcomes are missing and a complete case analysis is not valid, the primary analysis will be a joint Bayesian analysis of the missingness mechanism and outcome model. In general, we will not jointly model or impute missing outcomes. If outcomes are missing and a complete case analysis is not used, then we will report descriptive statistics for this outcome and the reasons that individuals were lost to follow up.

### Oversight and monitoring

Safety oversight will be under the direction of an independent DSMB composed of five individuals with expertise in pediatric emergency medicine, trial methodology, epidemiology and biostatistics operating under an approved charter. The DSMB will meet biannually and *ad hoc* as needed. They will receive a blinded interim report on safety but can, if necessary, ask for unblinding. The DSMB will provide its input to the Sponsor-Investigator and Steering Committee.

#### Adverse event reporting and harms

AEs will be defined as any untoward medical occurrence in a patient or clinical investigation subject administered a pharmaceutical product and which does not necessarily have a causal relationship with this treatment. All noxious and unintended responses to a medicinal product related to any dose will be considered adverse drug reactions (ADRs). AEs will be monitored for during the enrolment ED visit, on the 21-day medical record review, and on all follow-up surveys. Serious adverse events (SAEs) and serious adverse drug reactions (SADRs) will be defined as any untoward medical occurrence that (i) results in death, (2) is considered a life threatening event (patient at imminent risk of death), (3) results in a persistent or significant incapacity or substantial disruption of the ability to conduct normal life functions, (4) prolongs an existing hospitalization, or (5) is an important medical event that may not result in death, be life-threatening, or require hospitalization, but may be considered serious when, based upon appropriate medical judgment of the site QI, they may jeopardize the participant and may require medical or surgical intervention to prevent one of the outcomes listed in this definition. Participants with AEs will be followed until the event is resolved, or the site QI has deemed the participant stable or the condition chronic.

#### Monitoring

Monitoring for quality and regulatory compliance and participant safety will be performed in accordance with a Trial Monitoring Plan. In Canada, monitoring will be performed by the University of Alberta’s Quality Management in Clinical Research (QMCR) office. QMCR is an independent unit housed within the University of Alberta’s central administration. It is independent of the trial sponsor and sponsor-investigator. In New Zealand and Australia, monitoring of Australian sites and New Zealand sites (other than Perth Children’s Hospital) will be provided by the senior study coordinator in Australia (located at Perth Children’s Hospital). A monitor, independent of the site study team, will monitor the Perth Children’s Hospital site. Monitoring will occur at least once per year at each study site during the trial recruitment period.

### Trial management

The sponsor-investigator (ACP) will delegate a member of her research team as the project manager to manage the day-to-day operations of this trial. The site QI and site coordinator will be responsible for the day-to-day operations at their site and responsible for following the protocol. The sponsor-investigator, project manager, all site investigators, and site coordinators, as well as representatives from the monitoring group, data management group, methods group and network coordinating centre will meet monthly during recruitment phase. Data collection and entry is the responsibility of the clinical trial staff at the site under the supervision of the site QI. Site QIs will be responsible for ensuring the accuracy, completeness, and timeliness of data entries. The Women and Children’s Health Research Institute (WCHRI) at the University of Alberta (Edmonton) will act as the Data Coordinating Centre (DCC) housing all study data and providing data management services. Data will be entered into REDCap[60, 61], a validated electronic, web based, data capture system which is housed on secure servers at the University of Alberta. Internal quality checks within REDCap, such as automatic range checks, will be used to identify data that appear inconsistent, incomplete, or inaccurate. The Innovative Pediatric Clinical Trials (iPCT) Methods Group at the University of Toronto will be responsible for the data analysis.

This trial is part of a collaborative approach to pediatric clinical research in Canada. Under the KidsCan-PERC iPCT initiative four clinical trials will share resources and infrastructure with the iPT Coordinating Centre located at the University of Manitoba. The iPCT Steering Committee will be composed of the sponsor-investigators for all four trials, the iPCT PI (TPK) and coordinator, and other KidsCan-PERC iPCT initiative members. The iPCT Steering Committee will meet quarterly to review trial progress and network milestones.

### Confidentiality

All study related information will be securely stored at the study site in locked file cabinets in areas with limited access. The study participant’s contact information which will be linked to a study ID. This information will be securely stored at each site on a password protected document on a secure server. All records will be kept for as long a period as dictated by the reviewing REB, institutional policies, sponsor requirements or regulatory requirements. The study monitors, other authorized representatives of the sponsor, representatives of the REB, or regulatory agencies may inspect all documents and records required to be maintained by the site QI, including but not limited to, medical records and pharmacy records for the participants in this study. The study site will permit access to such records. Only de-identified study participant research data will be stored on the REDCap system housed by the DCC. The sponsor-investigator, the WCHRI DCC and the iPCT Methods group will be given access to the final trial data set. Site QIs will have direct access to their own site’s data set and will have access to other sites data by request. iPCT Network Steering Committee members will be given access to data by request. De-identified data from this study may be requested by other researchers 2 years after the publication of the primary manuscript by contacting the principal investigator (ACP). Consent for this data sharing will be obtained at the time of consent to enrolment in the trial. Researchers that wish to study the data must have the new study approved by a research ethics board and sign an agreement ensuring confidentiality and restricting data use only to the approved study. The study protocol, statistical analysis plan, and informed consent form will also be available on request.

### Knowledge Dissemination

Study results will be published in international scientific journals and will be presented at national and international conferences. The results of this study will also be communicated through press and social media. A formal authorship plan has been developed. plan for sharing the trial results with the caregivers of participating infants is in place.

### Protocol Amendments

The Sponsor-Investigator will communicate in writing all protocol modifications to the site QIs and site coordinators in writing and when necessary, undertake updated protocol training. All protocol modifications will also be submitted to the relevant REBs and regulatory authorities (e.g., Health Canada) as required by regulations. Any important modification that could affect enrolled participants will also be communicated to their caregivers.

### Post-trial care

No specific provisions have been made for ancillary and post-trial care or compensation for those who suffer harm from trial participation outside that usually provided by the jurisdictions in which the trial is undertaken.

### Summary of amendments to trial design and trial implementation in response to the SARS-Co-V-2 pandemic

The SARS-Co-V-2 pandemic resulted in challenges for clinical trials worldwide. For this trial, that was recruiting infants with respiratory illness in the ED these challenges were particularly significant. Study sites were affected by restrictions in the use of nebulizers to deliver inhaled medications in both clinical care and research due to concerns of the risk of respiratory virus transmission[62–65]. Due to public health restrictions, there was also a marked initial reduction in RSV circulating in the community and a resultant decrease in infants presenting with bronchiolitis. This reduction in RSV infections was then followed by alterations in the RSV seasonality[66–69]. The investigators for this study interpreted these extenuating circumstances related to the SARS-Co-V2 pandemic as a threat to the study’s feasibility[70]. In response to these challenges the investigators undertook two mitigation strategies. The first mitigation strategy was to move to allow sites to deliver the inhaled epinephrine by either nebulizer or by MDI-s depending on their site policy’s regarding nebulizer use (implemented December 2020, 155 participants enrolled). The second mitigation strategy was to have sites move to screen and enroll year-round to address the change in RSV seasonality. This change allowed sites to capture the variation in RSV season timing (implemented December 2021, 268 participants enrolled). In parallel to these pandemic related changes, we also reconsidered our frequentist analytic approach. We chose to move to a Bayesian approach for data analysis and inference due to increased acceptance of Bayesian approaches to clinical trial analysis[71] and availability of expertise (implemented December 2021, 268 participants recruited, 16.6% of original sample size). This resulted in a change in our sample size calculation for the trial from 1616 to 864. However, there were no changes to research question, the trial hypothesis, or primary or secondary outcomes.

## DISCUSSION

Bronchiolitis continues to exert a significant burden of illness worldwide. Given the known synergy between beta-agonists and steroids in asthma[12–15], the biological evidence for synergy[16, 17], and promising preliminary clinical results in bronchiolitis[8, 9], definitive evidence regarding the benefit of combined epinephrine and dexamethasone in bronchiolitis is needed to promote evidence-based practice for this common disorder. Further there are broad concerns about adverse effects by steroids by practicing clinicians[24] and given the potential risks of sustained high doses of dexamethasone examination of a shorter course of dexamethasone than studied[8] previously is essential.

In addition, the SARS-CoV-2 pandemic brought to the fore concerns that nebulized medication delivery may potentially increase the risk of respiratory virus transmission to health care staff through virus aerosolization[62–65]. While it was uncertain at the beginning of the pandemic as to whether SARS-CoV-2 would be a considerable cause of bronchiolitis, coronaviruses, as a group, were known to cause bronchiolitis[72–74]. Although health jurisdictions around the world have differing policies as to whether or not medication nebulization is an aerosolized generating medical procedure (AGMP)[75–80], the delivery of medication by metered dose inhaler with spacer (MDI-s) is considered a lower risk procedure and preferred by many jurisdictions and front line staff over nebulization[63–65]. Given these concerns, there is also a need to confirm specifically whether epinephrine delivered by MDI-s or by nebulizer is effective when combined with oral dexamethasone in reducing admissions for bronchiolitis.

Finally, given the heterogeneity across previous clinical trials, current guidelines, and clinical practice in defining bronchiolitis, we need to examine these interventions in a population which truly reflects patient care. The BIPED study will address these issues. The results will be highly generalizable internationally as recruitment will occur in three countries (Canada, Australia, and New Zealand). The patient-based outcomes assessed in this trial will be of wide interest to caregivers, clinicians, and healthcare administrators alike.

### Trial status

Study recruitment began 01 December 2018 and follow-up is expected to be complete by January 2025. As of 26 November 2024, 857 patients have been randomized. The current protocol is CTO 1423, dated 20 January 2023.

## Data Availability

De-identified data from this study may be requested by other researchers 2 years after the publication of the primary manuscript by contacting the principal investigator (ACP). Consent for this data sharing will be obtained at the time of consent to enrolment in the trial. Researchers that wish to study the data must have the new study approved by a research ethics board and sign an agreement ensuring confidentiality and restricting data use only to the approved study.

## Abbreviations

BIPED: Bronchiolitis in Infants – Placebo versus Epinephrine and Dexamethasone
ED: Emergency Department
PECARN: Pediatric Emergency Care Applied Research Network
CanBEST: Canadian Bronchiolitis Epinephrine Steroid Trial
PERC: Pediatric Emergency Research Network
MDI-s: metered dose inhaler with spacer
REB: Research Ethics Board
RSV: respiratory syncytial virus
SARS-CoV-2: severe acute respiratory syndrome coronavirus-2
RDAI: respiratory distress assessment index
QI: qualified investigator
REDCap: Research Electronic Data Capture
DSMB: Data Safety Monitoring Board
AE: adverse event
CRFs: (case report forms)
e-CRFs: electronic-case report forms
ICU: intensive care unit
CPAP: continuous positive airway pressure
ALC: average length criterion
ADR: adverse drug reaction
SAE: serious adverse event
SADR: serious adverse drug reaction
QMCR: Quality Management in Clinical Research
WCHR: Women and Children’s Health Research Institute
DCC: Data Coordinating Centre
iPCT: Innovative Pediatric Clinical Trials
AGMP: aerosolized generating medical procedure

## Patient and public involvement

Members of the CHEO Research Institute Patient and Family Advisory Committee reviewed the consent form and survey questions for clarity.

## Acknowledgements

The authors would like to acknowledge this is research has been facilitated by the WCHRI DCC through the generous support of the Stollery Children’s Hospital Foundation. Amphastar Pharmaceuticals^R^ (https://www.amphastar.com) provided Primatene Mist^R^ and placebo MDIs and Trudell Medical International^R^ (https://www.trudellmed.com/ca/en-ca) provided the spacers for use with the MDIs, free of charge. The following are members of the KidsCAN-PERC iPCT and PREDICT BIPED Study Team: Eleanor Pullenagyeum (Hospital for Sick Children Research Institute, Toronto, Ontario, Canada), Rick Watts (WCHRI, Edmonton, Alberta, Canada), and David Rios (Hospital for Sick Children Research Institute, Toronto, Ontario, Canada).

